# Evaluation of saliva sampling procedures for SARS-CoV-2 diagnostics reveals differential sensitivity and association with viral load

**DOI:** 10.1101/2020.10.06.20207902

**Authors:** Pieter Mestdagh, Michel Gillard, Marc Arbyn, Jean-Paul Pirnay, Jeroen Poels, Jan Hellemans, Eliana Peeters, Veronik Hutse, Celine Vermeiren, Maxime Boutier, Veerle De Wever, Patrick Soentjens, Sarah Djebara, Hugues Malonne, Emmanuel André, John Smeraglia, Jo Vandesompele

## Abstract

Nasopharyngeal sampling has been the preferential collection method for SARS-CoV-2 diagnostics. Alternative sampling procedures that are less invasive and do not require a healthcare professional would be more preferable for patients and health professionals. Saliva collection has been proposed as such a possible alternative sampling procedure. We evaluated the sensitivity of SARS-CoV-2 testing on two different saliva collection devices (spitting versus swabbing) compared to nasopharyngeal swabs in over 2500 individuals that were either symptomatic or had high-risk contacts with infected individuals. We observed an overall poor sensitivity in saliva for SARS-CoV-2 detection (30.8% and 22.4% for spitting and swabbing, respectively). However, when focusing on individuals with medium to high viral load, sensitivity increased substantially (97.0% and 76.7% for spitting and swabbing, respectively), irrespective of symptomatic status. Our results suggest that saliva cannot readily replace nasopharyngeal sampling for SARS-CoV-2 diagnostics but may enable identification of cases with medium to high viral loads.

## Introduction

Massive RT-qPCR based testing for the presence of SARS-CoV-2 RNA is a key element in the strategy to control the current COVID-19 pandemic. At present, collecting samples from the upper respiratory tract is recommended for diagnostic testing by the World Health Organization and (American and European) Centers for Disease Control and Prevention, with nasopharyngeal swabs being considered the standard collection procedure. While extremely sensitive, this sampling method is not patient-friendly and requires a healthcare professional with personal protective equipment to collect. Before the outbreak of SARS-CoV-2, several studies have reported on the utility of saliva for testing respiratory viruses^1–4^. The non-invasive nature of the specimen collection in a simple container, enabling home sampling, and the absence of swab and transport tube requirement (both plagued by global supply shortages), make saliva a very attractive biomaterial. While several recent studies have documented the potential utility of saliva for diagnostic testing of SARS-CoV-2^5–11^, these studies suffer from one or more limitations, i.e. non-paired study design, small cohorts, testing in biased populations such as previously confirmed positive cases and/or hospitalized patients. Here, we set out to prospectively evaluate the utility of saliva for diagnostic testing of SARS-CoV-2 using a large population of more than 2,500 individuals, including pauci-symptomatic and a-symptomatic participants, in triage centers in Belgium with matched nasopharyngeal and saliva collection at the same moment. We evaluated 2 types of saliva collection devices and all samples were analyzed by 2 independent laboratories.

## Results

### Study design

Study samples were collected at triage centers in Belgium by trained mobile teams. Individuals were sampled using 3 different procedures: (1) a nasopharyngeal swab sample representing the standard comparator for SARS-CoV-2 diagnostics, (2) a saliva sample collected through self-sampling with a commercial saliva spitting device (Norgen Biotek) and (3) a saliva sample collected through self-sampling with a commercial oral swabbing device (DNA Genotek) (Figure 1). Individuals were also asked to complete a survey enquiring about age group, ease of use of both devices, comfort of saliva versus nasopharyngeal sampling and (a)symptomatic status (Supplemental Table 1). All samples were analyzed in two independent test labs, applying different RNA extraction and RT-qPCR workflows (see Materials and Methods for details). Compatibility of both workflows with both saliva sampling devices (i.e. compatibility with the RNA preservation buffers in these devices) was validated using a 3-point 32-fold serial dilution of inactivated viral stock in saliva from healthy donors collected with both devices (Supplemental Figure 1).

**Figure 1.**
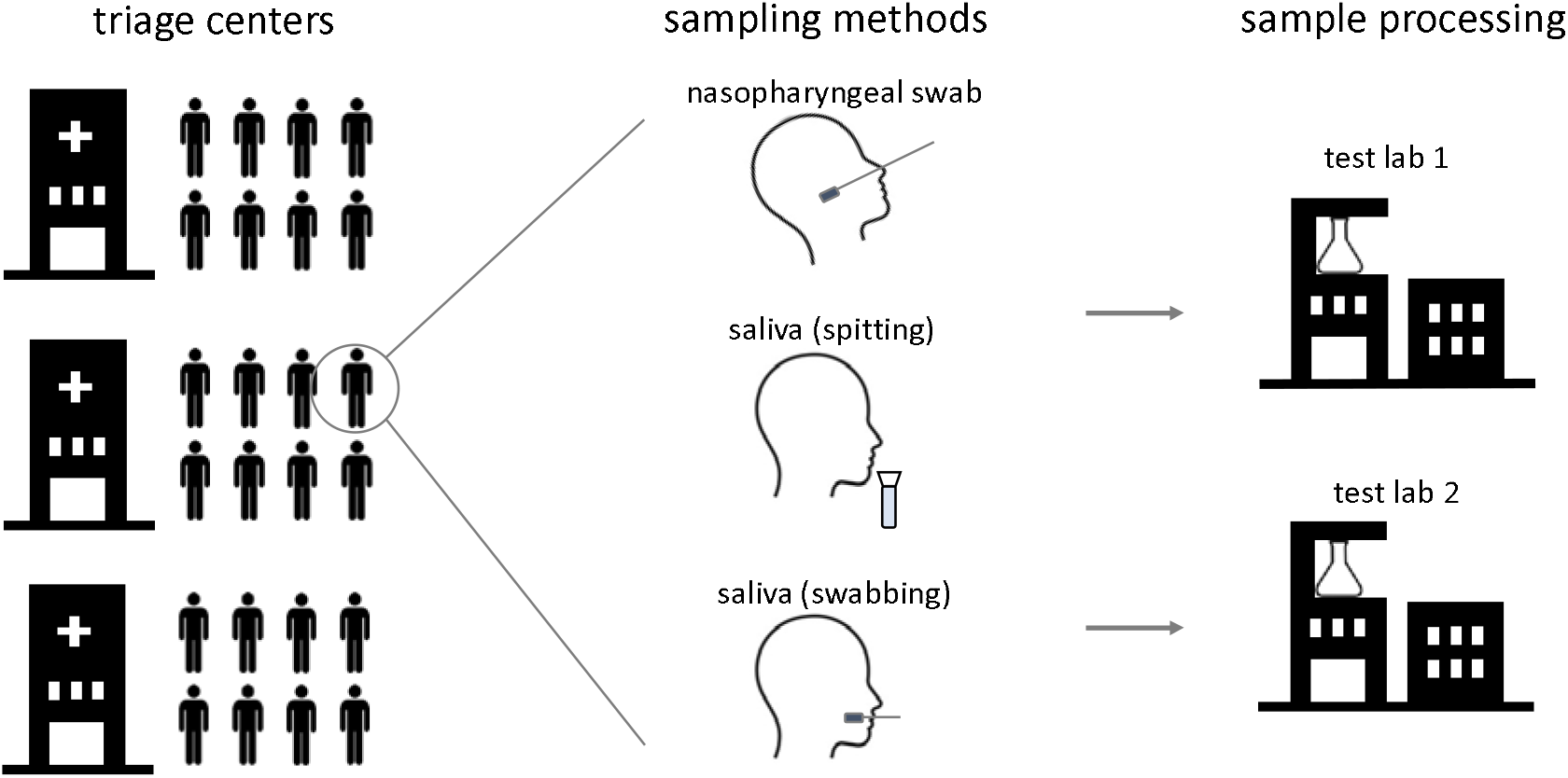
Overview of study design. Study participants were sampled at triage centers using a nasopharyngeal swab and two different saliva collection devices. Samples were processed at 2 different test labs using independent sample processing workflows.

The majority of study participants, 2289 individuals, was sampled with a nasopharyngeal swab and both saliva collection methods. Another 390 and 205 individuals were sampled with a nasopharyngeal swab and the saliva swabbing device or spitting device, respectively. While saliva sampling was generally perceived as more comfortable than nasopharyngeal sampling (Supplemental Figure 2A), study participants scored the ease of use of the saliva swabbing device significantly higher than that of the saliva spitting device (p < 10^−10^, Mann-Whitney test, Supplemental Figure 2B).

**Figure 2.**
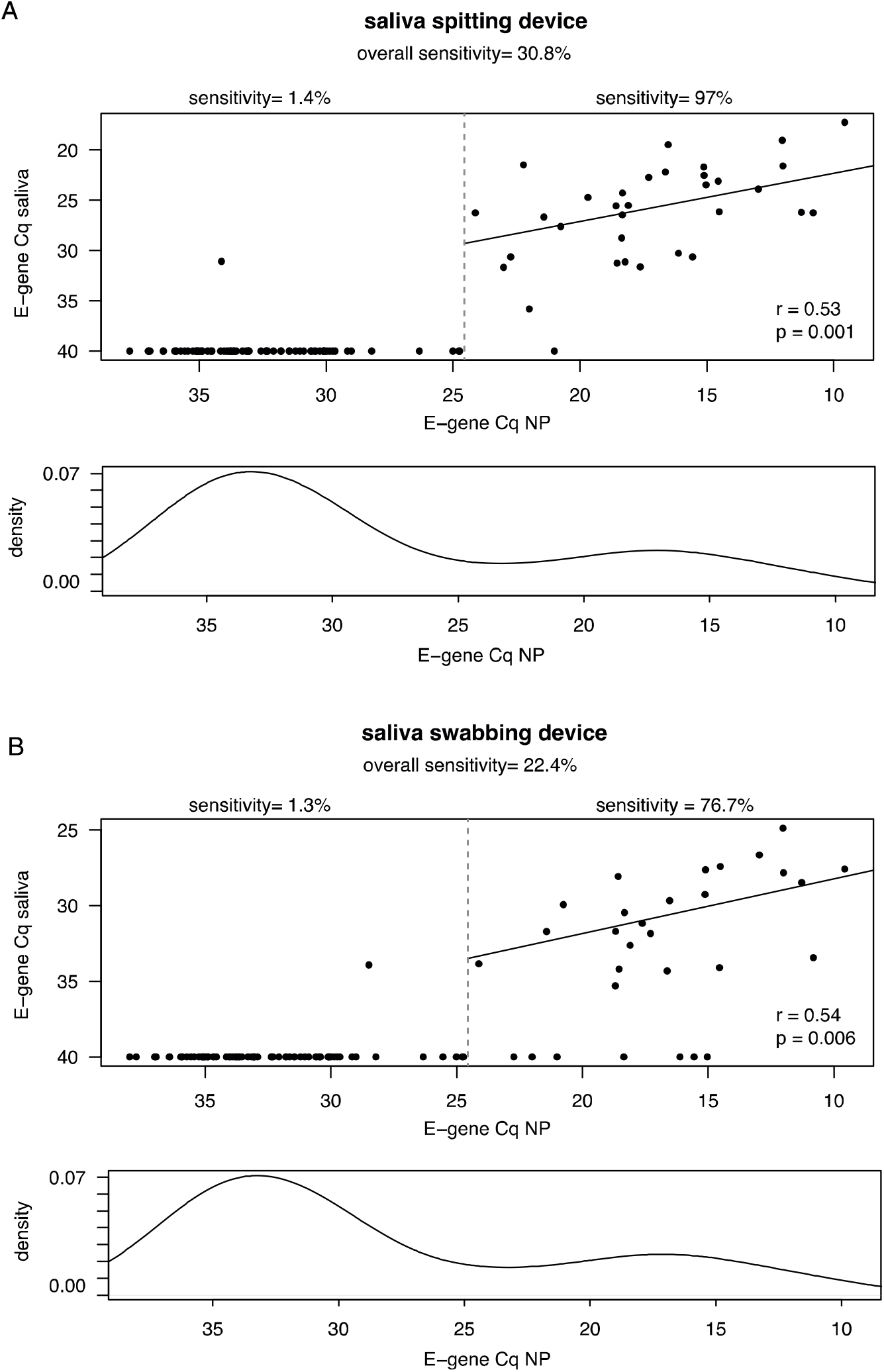
E-gene Cq values in SARS-CoV-2 positive nasopharyngeal samples and matching saliva samples. (A) Results obtained with the saliva spitting device. (B) Results obtained with the saliva swabbing device. Plots indicate the coefficient of determination (R^2^) value and p-value (Pearson’s correlation test) calculated using only those samples with a nasopharyngeal E-gene Cq-value < 24.5.

### Saliva sampling identifies individuals with medium to high viral load

Out of 2884 nasopharyngeal swab samples analyzed by test lab 1, 117 (4.0%) were SARS-CoV-2 positive. There were 107/117 nasopharyngeal positive samples for which a matching saliva spitting sample was available, and 107/117 nasopharyngeal positive samples for which a matching saliva swabbing sample was available. We observed 33/107 (sensitivity = 30.8%; CI= 22.5%-40.6%) saliva spitting samples and 24/107 (sensitivity = 22.4%; CI=15.2%-31.7%) saliva swabbing samples that were SARS-CoV-2 positive, indicating reduced overall sensitivity in saliva for SARS-CoV-2 detection (Figure 2). However, we observed a significantly higher nasopharyngeal viral load in patients with a true-positive saliva sample compared to patients with a false-negative saliva sample (spitting device: log_2_FC = 14.89, p=3.79 x 10^−15^; swabbing device: log_2_FC = 14.7, p=3.67 x 10^−12^, Mann-Whitney test.) (Supplemental Figure 3). Individuals with an E-gene Cq > 24.5 in the nasopharyngeal sample (corresponding to 56 000 copies/ml viral transport medium as determined by digital PCR and further referred to as low viral load, see Materials and Methods) almost always presented with a negative saliva sample (sensitivity = 1.4%; CI=0.07%-8.3% and sensitivity = 1.3%; CI=0.07%-8.0% in the saliva spitting and saliva swabbing sample, respectively) (Figure 2). In contrast, for individuals with a high viral load (E-gene Cq < 24.5 in the nasopharyngeal sample), concordance between the nasopharyngeal and matching saliva sample improved dramatically, especially for the saliva spitting device resulting in high sensitivity in this subgroup (sensitivity = 97.0%; CI=82.4%-99.8% and sensitivity = 76.7%; CI=57.3%-89.4% for the saliva obtained by spitting and swabbing, respectively) (Figure 2). In addition, we observed a significant positive correlation between E-gene Cq-values in the nasopharyngeal and saliva samples for those individuals with high viral load (spitting device: r = 0.53, p = 0.001; swabbing device: r = 0.54, p = 0.006; Pearson). Notably, similar findings were observed in test lab 2 (Supplemental Figure 4).

**Figure 3.**
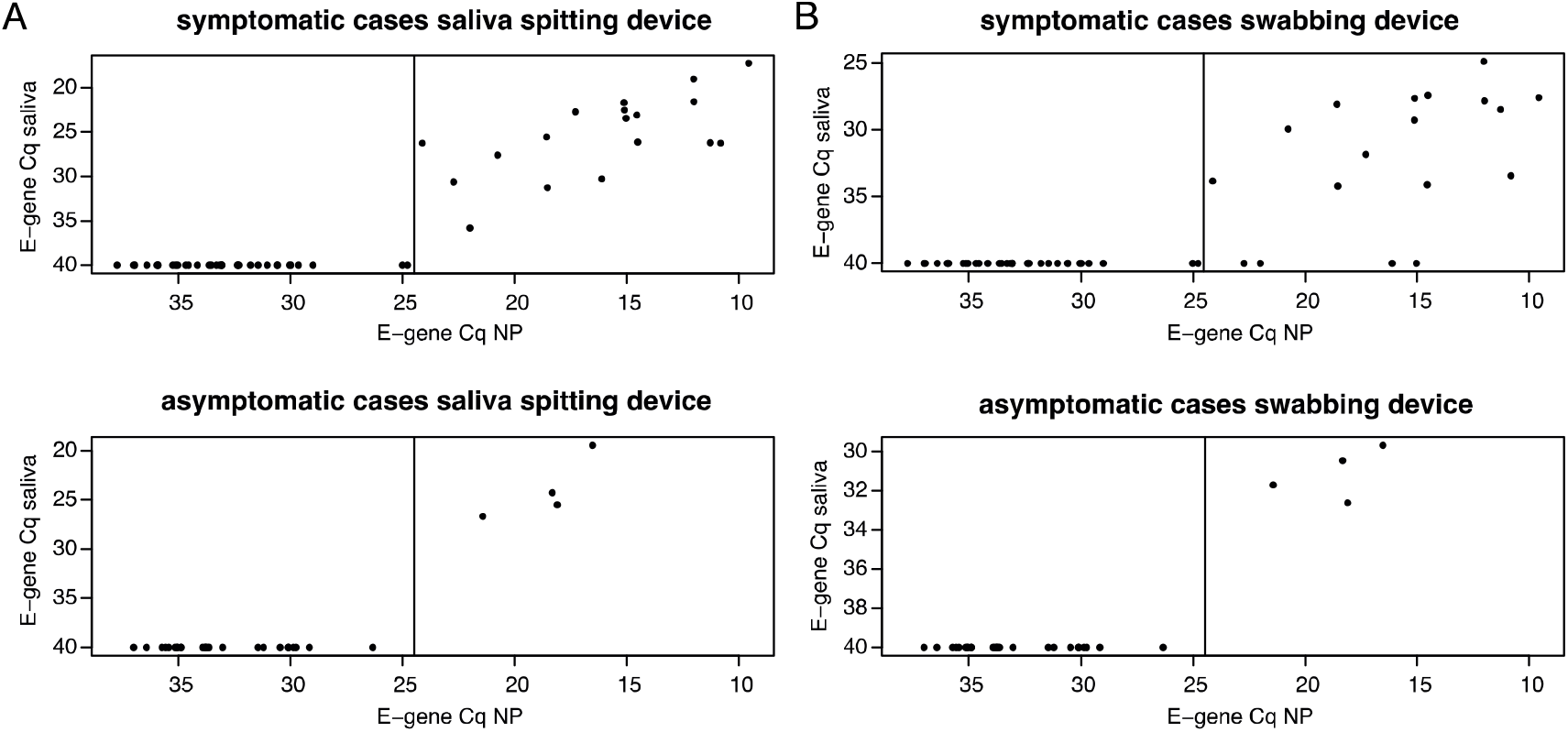
E-gene Cq values in nasopharyngeal positive samples and matching saliva samples from symptomatic and asymptomatic cases. (A) Results obtained with the saliva spitting device. (B) Results obtained with the saliva swabbing device.

To assess whether the poor sensitivity in saliva for SARS-CoV-2 detection could be due to sampling issues during saliva collection, we quantified the human gene RPS18 using RT-qPCR on a representative set of RNA samples used for SARS-CoV-2 detection. No differences in RPS18 levels were observed between saliva positive and negative samples for any of the saliva sampling devices, suggesting that sampling issues do not explain the false negative saliva samples (Supplemental Figure 5A-B).

We then evaluated whether there were study participants for whom the nasopharyngeal sample was negative and at least one of the saliva samples positive. Test lab 1 identified 12 such cases while test lab 2 identified 8 cases (Supplemental Table 2 and 3). These cases were mainly found among the saliva spitting samples (FPR, defined as the % saliva-positive that are negative on the NP specimen = 26.7%; CI=15.1%-42.2% and FPR = 7.7%; CI=1.3%-26.6% for saliva spitting and swabbing samples, respectively) and typically presented with very low viral loads (mean E-gene Cq saliva spitting samples = 32.6). Of note, only 2 cases were commonly identified by both test labs and these two cases were the only ones that were positive for both the spitting and swabbing saliva device. To assess whether the negative result in the nasopharyngeal sample was due to sampling issues, we also quantified RPS18 on a subset of these samples. Out of 4 SARS-CoV-2 negative nasopharyngeal samples (that had a matching positive saliva sample), 1 showed extremely low levels of RPS18 (Cq > 35) while the remaining 3 had RPS18 Cq-values similar to those of nasopharyngeal positive samples (Supplemental Figure 5C). These results suggest that nasopharyngeal sampling issues may explain some but not all of these cases.

### SARS-CoV-2 detection in saliva is dependent of symptomatic status

For 2172 study participants, we were able to register presence or absence of symptoms for COVID-19 (see Materials and Methods for clinical definition). From these, 1412 (65.0%) were symptomatic, 705 (32.5%) were asymptomatic and 55 (2.5%) indicated they experienced symptoms in the 2 weeks preceding the test. The latter group was excluded from further analyses because of the limited number of subjects.

The fraction of subjects that were SARS-CoV-2 positive in the nasopharyngeal sample was similar in the symptomatic and asymptomatic group, 4.67% and 4.96% respectively, however the symptomatic group was enriched for cases with high viral load (E-gene Cq <24.5, p = 0.042, Fisher’s exact test). As a result, sensitivity in saliva for SARS-CoV-2 detection was higher among symptomatic cases (sensitivity = 34.6%; CI=22.3%-49.2% and sensitivity = 26.9%; CI=16.0%-41.3% for spitting and swabbing saliva device respectively) compared to asymptomatic cases (sensitivity = 13.3%; CI=4.4%-31.6% for both the spitting and swabbing saliva device) (Figure 3). Nevertheless, among subjects with high viral load in the nasopharyngeal sample, sensitivity in saliva was high, irrespective of symptomatic status (saliva spitting device: sensitivity = 100%; CI=78.1%-100% in symptomatic cases, sensitivity = 100%; CI=46.3%-100% in asymptomatic cases; saliva swabbing device: sensitivity = 77.8%; CI=51.9%-92.6% in symptomatic cases, sensitivity = 100%; CI=39.6%-100% in asymptomatic cases).

## Discussion

Saliva sampling for COVID-19 diagnostics has been put forward as an alternative for nasopharyngeal sampling in a number of independent studies. A rapid review of the literature estimated a pooled sensitivity of SARS-CoV-2 testing on saliva versus nasopharyngeal samples as high as 97%^12^. Here, we compared the sensitivity of two different saliva collection devices to the nasopharyngeal swab in over 2500 individuals that were sampled at different triage centers in Belgium. In contrast to the current literature, we observed a substantially lower SARS-CoV-2 test positivity rate in saliva than in nasopharyngeal samples. When focusing on individuals with a high viral load (more than 56 000 copies/ml viral transport medium), concordance improved dramatically, especially in saliva samples produced through spitting. There are several potential reasons for the discrepancy between results presented here and current literature reports. First, the study population may be very different. Most of the studies reported in literature predominantly include symptomatic patients (whether or not hospitalized) that are more likely to have a high viral load. Our study included individuals visiting triage centers to get a diagnostic test, either because they presented with COVID-19 symptoms or because they had been in contact with an infected individual. These individuals were not critically ill and often did not have symptoms. Secondly, while our study compared two different devices for saliva collection, we cannot exclude the possibility that these devices are less suitable for saliva collection compared to what has been used in other studies. Of note, we observed a differential sensitivity between both devices and conclude that saliva sampling through spitting is more sensitive than swabbing. Whether the order of saliva collection (participants were asked to first swab, then spit) could impact these results remains to be investigated. Finally, other co-variates, including time of sampling and stage of infection at sampling may also impact results. Additional studies are required in order to further investigate the impact of these factors.

The conclusions presented in this study are based on results generated by 2 different test labs. While results from both labs independently support these conclusions, we also observed some discrepancies between both labs in the test results from individual samples (i.e. samples that were called positive by lab 1 and negative by lab 2 or vice versa). These discrepancies were primarily observed in samples with low viral load and may thus, at least in part, be explained by sampling noise. Intrinsic differences in SARS-CoV-2 quantification between platforms (i.e. E-gene versus S-gene, N-gene and ORF1ab-gene) as well as occasional false positives could also play a role. We note that discrepant samples were observed throughout the entire duration of the study and therefore do not result from a single problematic test batch.

Our study suggests that saliva sampling cannot replace the standard nasopharyngeal swab for COVID-19 diagnostics. Nevertheless, because of its ease of use and compatibility with self-sampling, saliva sampling could play a role in systematic screening campaigns that aim to identify asymptomatic cases with medium to high viral loads. However, based on results presented here, such screening campaigns would fail to identify low positives. Whether, and to what extent, these low positives are capable of spreading the virus requires further investigation.

## Materials and Methods

### Patient samples

Nasopharyngeal swabs were taken by a healthcare professional as a diagnostic test for SARS-CoV-2, as part of the Belgian national testing platform. The individuals were tested in triage centers, between June 2020 and July 2020. In parallel, saliva samples were collected using Norgen Biotek’s Saliva RNA Collection and Preservation Device Dx #53800 (for collection of 2 ml of saliva through spitting), DNA Genotek’s ORAcollect RNA device # ORE-100 (for saliva collection through swabbing), or both. Participants in the study were asked not to eat, drink, smoke or use chewing gum in the last 30 minutes preceding saliva sampling. After sample collection, a short survey was completed to enquire about age group, ease of use of both saliva devices, comfort of saliva sampling versus nasopharyngeal sampling and symptomatic status. To enquire about symptomatic status, we applied the case definition of Sciensano, the Belgian Institute for Health. This study S64125 was approved by the ethical review committee of the University Hospital of Leuven on May, 29 2020.

### Power calculation

The sample size was computed for assessment of a hypothesis on non-inferior SARS-CoV-2 positivity on saliva compared to on nasopharyngeal specimen in paired testing as proposed by Tang^13^, using target values from a systematic review^12^. We accepted a confidence of 95%, a power of 80%, a sensitivity of the test in NP samples of 95%, a proportion of saliva-/NP+ samples 5% and 0.90 as benchmark for the relative positivity rate (saliva/NP), which yielded 84 SARS-CoV-2+ subjects needed. These could be found in a study population of 841 to 8410 subjects assuming a prevalence of 1% to 10%. Given the substantially larger contrast in test positivity between saliva and NP specimens, study enrolment was stopped when reaching 2884 inclusions.

### SARS-CoV-2 RT-qPCR test lab 1

RNA extraction was performed using the Total RNA Purification Kit (Norgen Biotek #24300) according to the manufacturer’s instructions using 200 µl viral transport medium (for the nasopharyngeal swab, i.e. Zymo Research’s DNA/RNA Shield # R1100-250) or 200 µl saliva, 200 µl lysis buffer and 200 µl ethanol, with processing using a centrifuge (5810R with rotor A-4-81, both from Eppendorf). RNA was eluted from the plates using 50 µl elution buffer (nuclease-free water), resulting in approximately 45 µl eluate. RNA extractions were simultaneously performed for 94 patient samples and 2 negative controls (nuclease-free water). After addition of the lysis buffer, 4 µl of a proprietary 700 nucleotides spike-in control RNA (5000 copies) and carrier RNA (200 ng of yeast tRNA (Roche #10109517001) was added to all 96 wells from the plate). To the eluate of one of the negative control wells, 7500 RNA copies of positive control RNA (Synthetic SARS-CoV-2 RNA Control 2, Twist Biosciences #102024) were added.

Six µl of RNA eluate was used as input for a 20 µl duplex RT-qPCR reaction in a CFX384 qPCR instrument using 10 µl iTaq one-step RT-qPCR mastermix (Bio-Rad #1725141) according to the manufacturer’s instructions, using 250 nM final concentration of primers and 400 nM of hydrolysis probe. Primers and probes were synthesized by Integrated DNA Technologies using clean-room GMP production. For detection of the SARS-CoV-2 virus, the Charité E gene assay was used (FAM)^14^; for the internal control, a proprietary hydrolysis probe assay (HEX) was used. Cq values were generated using the FastFinder software v3.300.5 (UgenTec). Only batches were approved with a clean negative control and a positive control in the expected range. Proper RNA extraction and RT-qPCR was confirmed by observing spike-in RNA signal in each sample well in the expected range.

### SARS-CoV-2 RT-qPCR test lab 2

RNA extraction was performed using a magnetic bead-based RNA extraction method developed by University of Liège (CoRNA kit) and according to the recommended protocol. For nasopharyngeal swab samples, 200 µl were transferred to a proteinase K solution. For both saliva devices, 100 µl of saliva was transferred to a lysis buffer mix (proteinase K solution + lysis buffer). The RNA extraction procedure was performed on samples spiked with the MS2 phage as internal control, and in presence of carrier RNA to increase RNA extraction efficiency. The multiplex RT-qPCR was performed on 5 µl of RNA eluate using TaqPath COVID-19 Combo Kit (comprising ORF1ab, N gene, S gene, and MS2 as internal control, #A47814, Applied Biosystems) and TaqPath 1-Step Multiplex Master Mix (no ROX) (#A28523, Applied Biosystems), following the manufacturer’s instructions. Cq values were generated using the FastFinder software v3.300.5 (UgenTec). Results were approved when a clean negative control and a positive control in the expected range were obtained. Correct RNA extraction and RT-qPCR setup was also confirmed by controlling MS2 amplification in each sample well (Cq <33).

### Digital PCR quantification of positive control RNA

In order to estimate the viral load concentration in nasopharyngeal transport medium equivalent with the threshold above which concordance between saliva and the nasopharyngeal is high, we used digital PCR to quantify the positive control RNA that was run in each batch of RNA samples quantified with RT-qPCR. This calibration point was then used to convert the threshold into copies per ml nasopharyngeal transport medium. Digital PCR was done on a QX200 instrument (Bio-Rad) using the One-Step RT-ddPCR Advanced Kit for Probes (Bio-Rad #1864022) according to the manufacturer’s instructions. Briefly, 22 µl pre-reactions were prepared consisted of 5 µl 4x supermix, 2 µl reverse transcriptase, 6 µl positive control RNA (125 RNA copies/µl), 15 mM dithiothreitol, 900 nM of each forward and reverse primer and 250 nM E-gene hydrolysis probe (FAM) (see higher). 20 µl of the pre-reaction was used for droplet generation using the QX200 Droplet Generator, followed by careful transfer to a 96-well PCR plate for thermocycling: 60 min 46 °C reverse transcription, 10 min 95 °C enzyme activation, 40 cycles of 30 sec denaturation at 95 °C and 1 min annealing/extension at 59 °C, and finally 10 min 98 °C enzyme deactivation. Droplets were analyzed by the QX200 Droplet Reader and QuantaSoft software. With an RNA input of 750 copies per reaction, the digital PCR result was 150 cDNA copies (or 20% of the expected number, a fraction confirmed by Dr. Jim Huggett for particular lot numbers of #102024, personal communication).

Based on the median Cq value of the positive control RNA of 24.55 for 1500 digital PCR calibrated cDNA molecules as input, the observed low viral load Cq cut-off value of 24.5 corresponds to 1500 copies per 6 µl RNA input. With an effective RNA elution volume of 45 µl and 200 µl transport medium used for RNA extraction, the low viral load Cq cut-off value of 24.5 in test lab 1 roughly corresponds to 56 000 copies/ml viral transport medium.

### Analytical platform compatibility testing

To validate compatibility of the workflows applied by test lab 1 and test lab 2, we created a saliva validation panel by collecting saliva from 3 different healthy donors using both saliva sampling devices. Donors did not eat or drink, smoke or use chewing gum for 30 minutes prior to saliva collection. For each collection device, saliva samples from all three donors were pooled and spiked with inactivated SARS-CoV-2 virus stock. More specifically, 50 µl of a 2.4-fold, 76.8-fold and 2457.6-fold dilution was added to 1950 µl of pooled saliva. A negative control saliva sample was generated by adding 50 µl of water. Saliva samples were aliquoted and processed by both test labs.

## Data Availability

All data are available as supplemental tables.

## Acknowledgements

The authors want to address special thanks to the doctors, the nurses and the paramedics of the Belgian Armed Forces and other teams involved in the massive sampling campaigns in the different triage centers. The authors want to thank Pierre Wattiau (GSK Biologicals) and Olivier Vandeputte (GSK Biologicals) for constructive discussions on study design and results and Els Dequeker (KU Leuven) for supervising the ethical approval and processing of the survey forms.

---

**Supplemental Figure 1.**
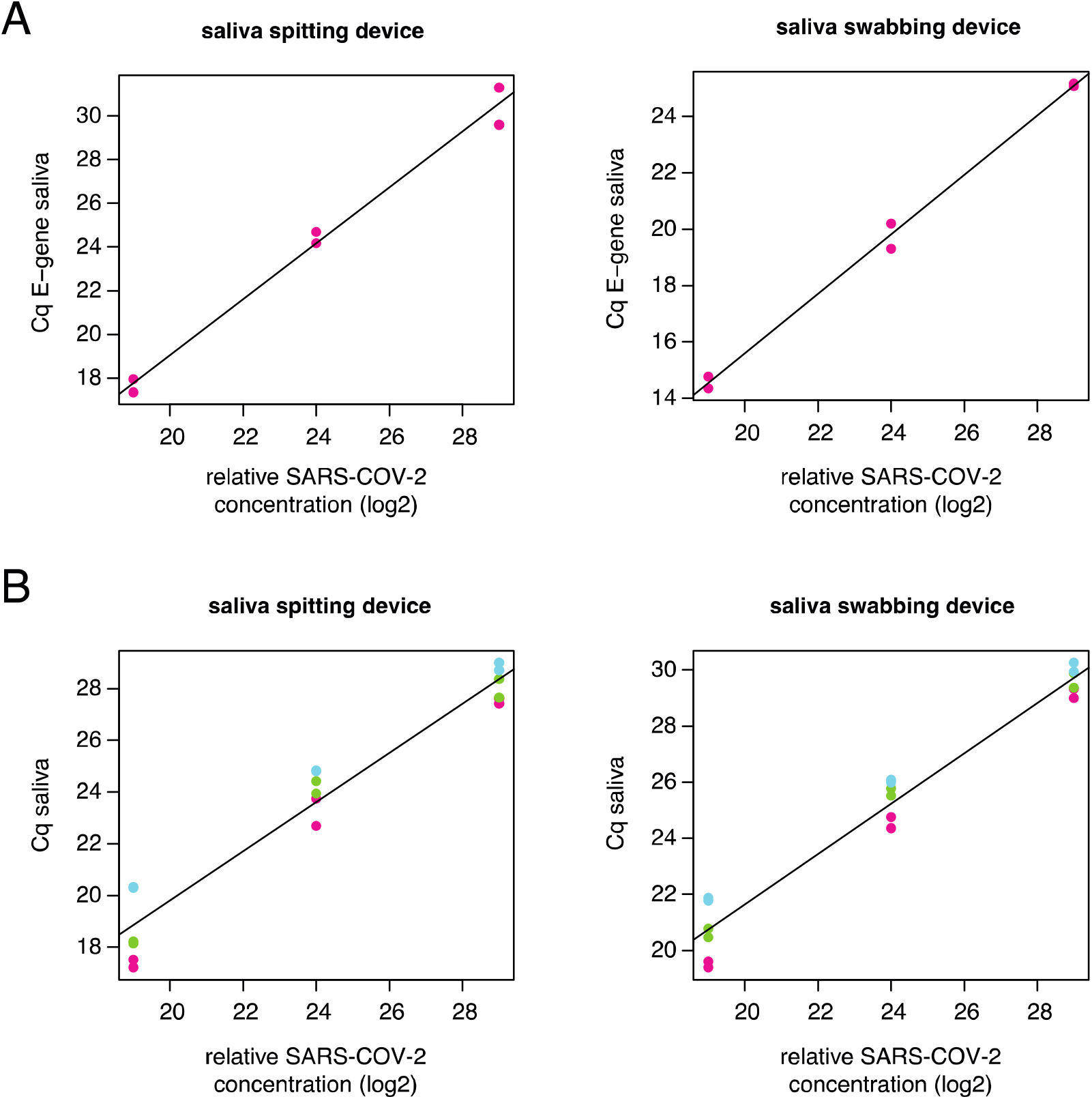
(A) E-gene Cq value quantified by test lab 1 on a pool of saliva from healthy donors spiked with inactivated SARS-CoV-2 at three different concentrations, for saliva collection by spitting (at left) and for swabbing (at right). (B) Cq-value of ORF1AB (pink), N-gene (green) and S-gene (blue) quantified by test lab 2 on a pool of saliva from healthy donors spiked with inactivated SARS-CoV-2 at three different concentrations.

**Supplemental Figure 2.**
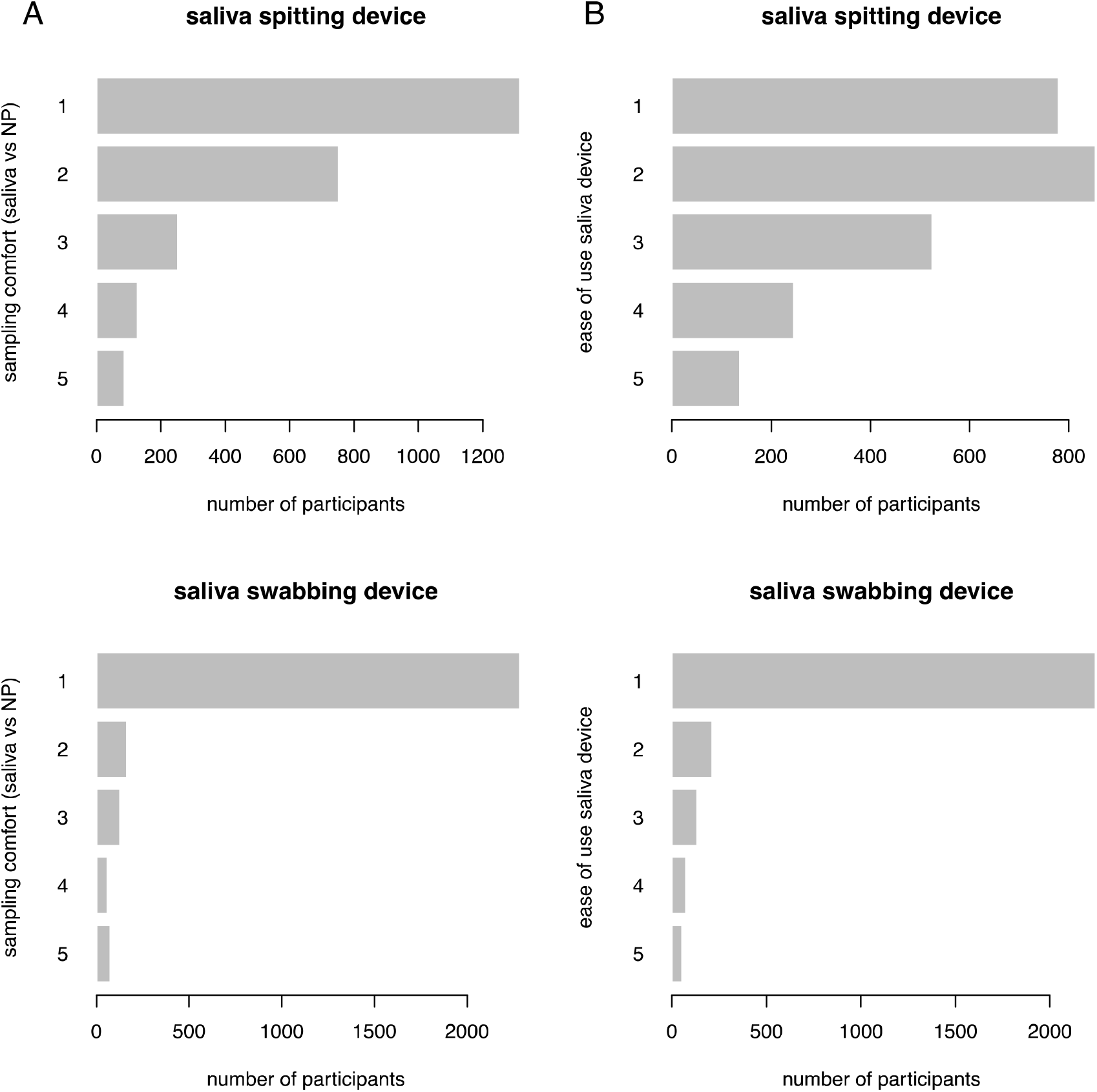
(A) Saliva versus nasopharyngeal (NP) sampling comfort, scored by study participants on a scale from 1 (more comfortable) to 5 (less comfortable) for both saliva sampling devices. (B) Ease of use of both saliva sampling devices, scored by study participants on a scale of 1 (easy to use) to 5 (difficult to use).

**Supplemental Figure 3.**
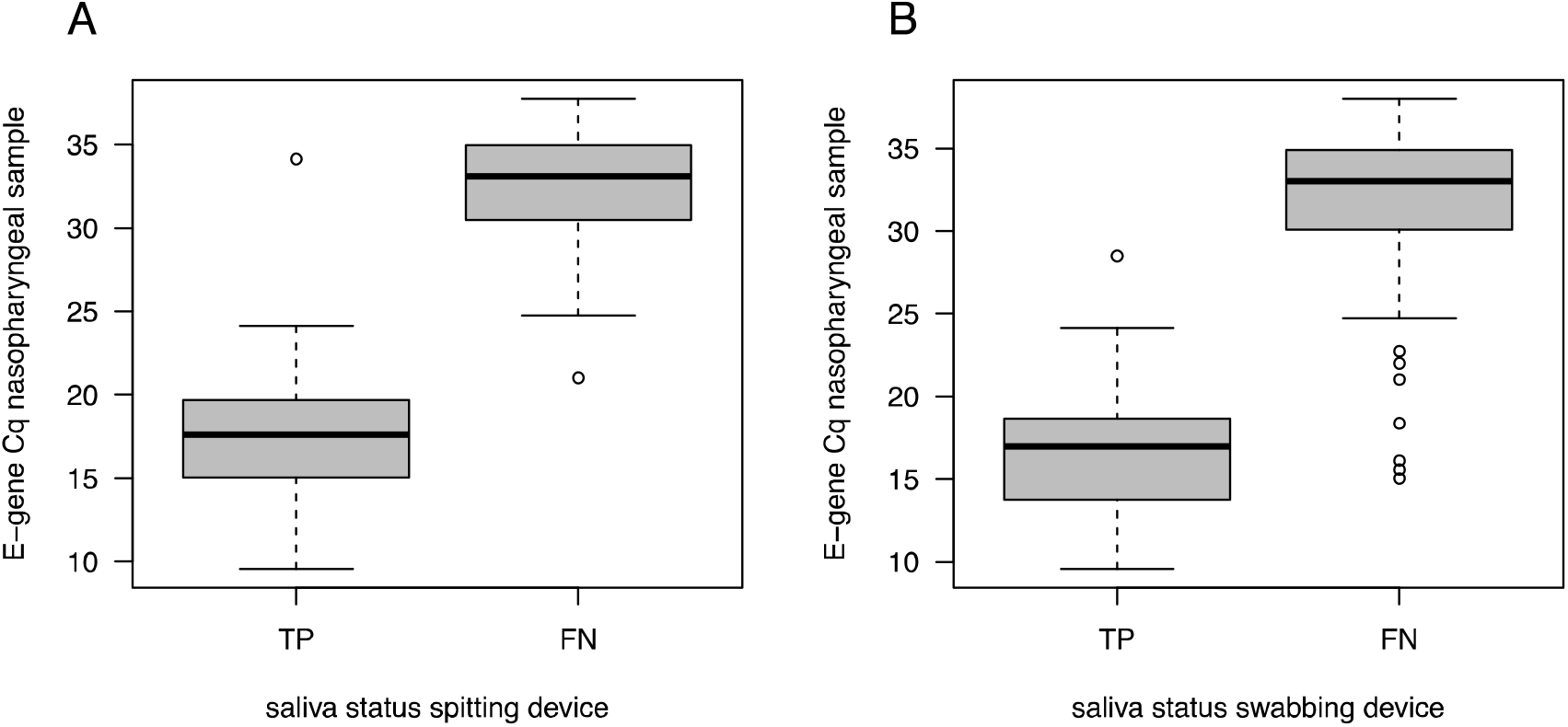
(A) E-gene Cq-value in nasopharyngeal samples for which the matching saliva spitting sample is a true positive (TP) or false negative (FN). (B) E-gene Cq-value in nasopharyngeal samples for which the matching saliva swabbing sample is a true positive (TP) or false negative (FN).

**Supplemental Figure 4.**
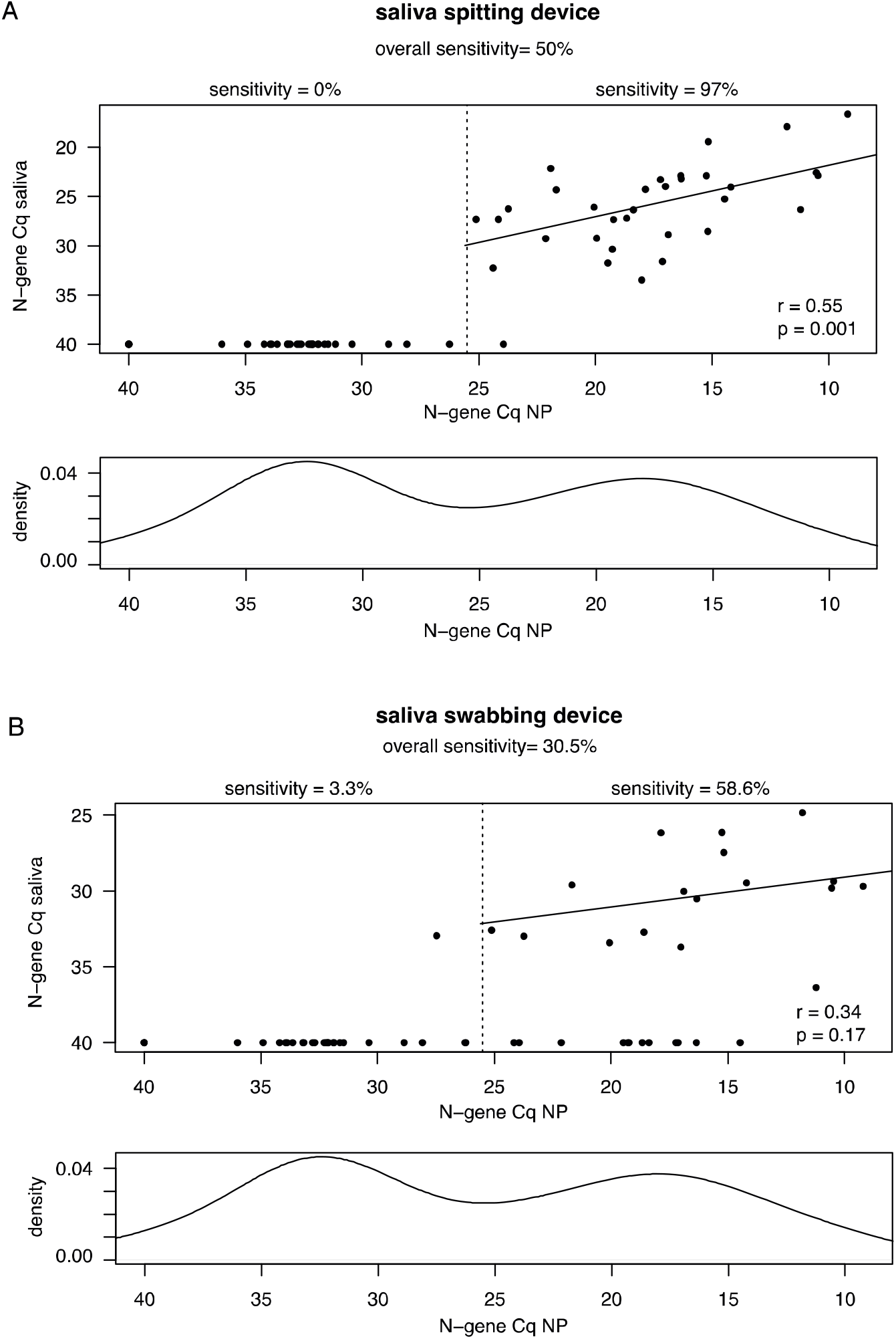
N-gene Cq values quantified by test lab 2 in nasopharyngeal positive samples and matching saliva samples. (A) Results obtained with the saliva spitting device. (B) Results obtained with the saliva swabbing device. Plots indicate the coefficient of determination (R^2^) value and p-value (Pearson’s correlation test) calculated using only those samples with a nasopharyngeal E-gene Cq-value < 25.5.

**Supplemental Figure 5.**
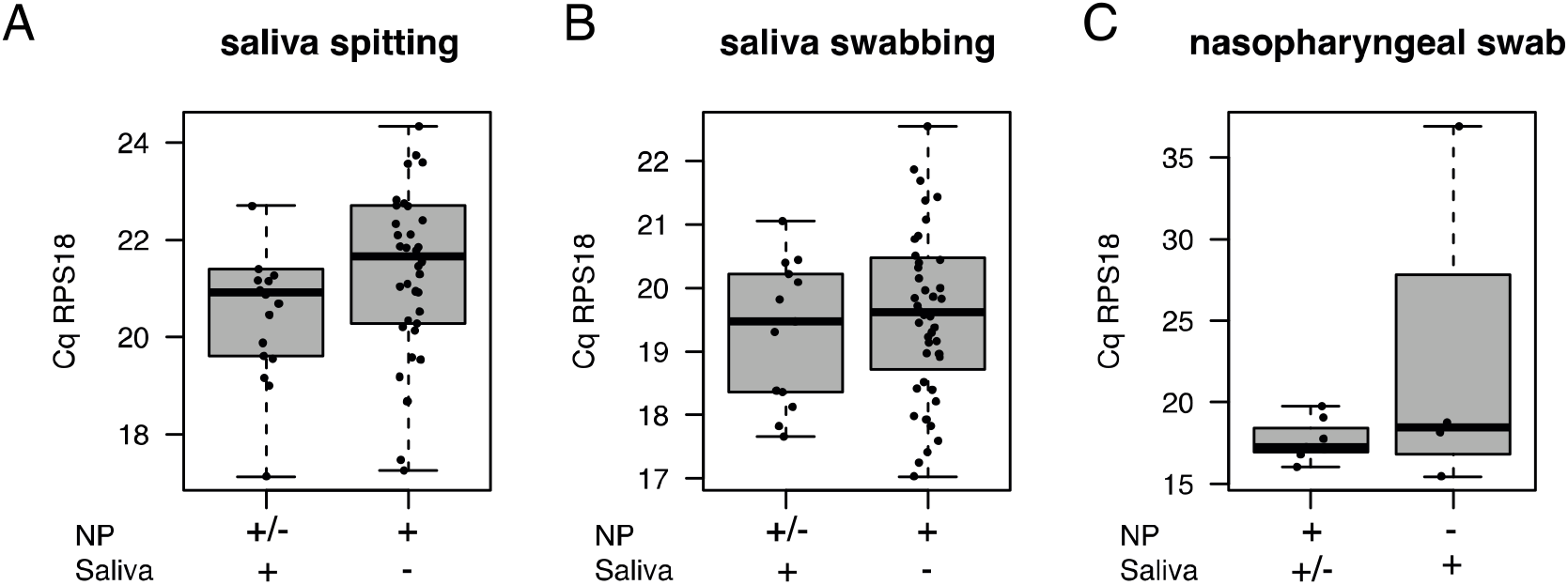
Abundance levels of RPS18 in SARS-CoV-2 positive and negative saliva samples (A,B). (C) Abundance levels of RPS18 SARS-CoV-2 positive and negative nasopharyngeal samples.

## Notes

### Competing Interest Statement

The authors have declared no competing interest.

### Funding Statement

No external funding was received

### Author Declarations

This study S64125 was approved by the ethical review committee of the University Hospital of Leuven on May, 29 2020.

